# Multimodal neuroimaging correlates of physical-cognitive covariation in Chilean adolescents. The Cogni-Action Project

**DOI:** 10.1101/2022.03.28.22273069

**Authors:** Carlos Cristi-Montero, Heidi Johansen-Berg, Piergiorgio Salvan

## Abstract

Health-related behaviours have been related to brain structural features; however, most literature in this domain comes from developed countries. In developing settings, such as Latin America, high social inequality is associated inversely with several health-related behaviours affecting brain development. Understanding the relationship between health behaviours and brain structure in such settings is particularly important during adolescence when key habits are acquired and ingrained. In this cross-sectional study, we carry out a multimodal analysis identifying a brain region associated with health-related behaviours (i.e., fatness, fitness, sleep problems and others) and cognitive/academic performance independent of socioeconomic status in a large sample of Chilean adolescents. Our findings suggest that the relationship between health behaviours and cognitive/academic performance involves a particular brain phenotype that could play a mediator role. These findings raise the possibility of promoting healthy behaviours in adolescence as a means to influence brain structure and thereby cognitive/academic achievement, independently of socioeconomic factors.

## Introduction

Behaviours emerge in response to particular situations, experiences, or repetitive stimuli which modulate our brain through an adaptive process ^1^. Although this process occurs throughout the lifespan ^2^, adolescence is key, as it is a critical period both for acquiring and ingraining habits and for extensive neural reorganisation ^2–4^.

Adolescents’ physical activity, physical fitness, diet, sleep and other lifestyle factors have been extensively studied in relation to health outcomes ^5,6^. However, beyond physical health outcomes, cognitive performance and academic achievement have been recently declared critical outcomes for children and adolescents by the World Health Organisation ^7^. In this sense, health-related behaviours are crucial during childhood due to their impact on brain features (i.e., structure, function, perfusion), hippocampal neurogenesis, neurotrophic factors release, cognition, learning abilities, and academic achievements ^5,6,8–12^. However, several studies and reports show a low rate of adolescents meeting the current physical activity and sleep recommendation, a high prevalence of obesity, and a poor healthy diet ^4,11,13,14^; thus creating a potential risk for healthy brain and cognitive development.

In addition, socioeconomic factors also play a critical relevant role not just in cognition and academic achievement, where greater deprivation is associated with worse outcomes, but also in brain development ^15,16^. Nonetheless, health-related behaviours such as physical fitness and physical activity could be leveraged as behavioural drivers to mitigate long-term consequences of early life adversity (i.e., poverty) on brain health ^16,17^. In this sense, developing countries and nations with high heterogeneity in income distribution present a significant challenge to reducing socioeconomic inequalities in health ^18,19^.

Although social determinants could modulate health-related behaviours and, in turn, brain, cognitive, and academic achievements in adolescents ^17,20,21^, most literature in this area comes from developed countries, being practically non-existent in children and adolescents living in Latin America ^22^. Chile is a particularly interesting case to study due to it progressively transitioning from low-to-middle to being a high-income country whilst retaining high inequality of income distribution (44.4%). Furthermore, its educational system segregates students into public, subsidised or private schools according to household income ^23^. Therefore, understanding whether health-related behaviours (i.e., fitness, physical activity and, more in general, healthy-lifestyles) relate to brain and cognitive health in adolescence, even in the presence of high heterogeneity in socioeconomic factors, is key to identifying modifiable lifestyle factors capable of improving developmental outcomes in the transition to adulthood.

Human lifestyles are complex, with multiple factors interacting and affecting each other simultaneously. Hence studying the association between individual variables risks missing the numerous interactions among them ^24,25^. Previous studies using multivariate statistical approaches have shown a heterogeneous contribution of each lifestyle factor with brain health indicators in adolescents (i.e., cognitive performance and mental health) ^20,26^ and diffuse brain-wide correlates (e.g., mainly structural and microstructural MRI metrics) ^27^.

Building on this work, here we tested the hypothesis that in a large sample of Chilean adolescents, healthy-lifestyle behaviours (i.e., fatness, fitness, sleep problems and others) were related to individual differences in cognitive skills and academic achievement (i.e., working memory, attention, maths, language), independent of socioeconomic factors, schools, and other confounds of no interest. In a sub-sample with neuroimaging, we also hypothesised that such behaviour–cognitive covariation may have structural and microstructural brain correlates and that these brain correlates may mediate the covariation between health-related behaviours and cognitive/academic achievements, independent of socioeconomic background.

## Methods

This cross-sectional study is part of the Cogni-Action Project ^28^ carried out from March 2017 to October 2019. The project was conducted according to the guidelines of the Declaration of Helsinki and approved by the Bioethics and Biosafety Committee of the Pontificia Universidad Católica de Valparaíso (BIOEPUCV-H103–2016) and was retrospectively registered (8/July/2020) in the Research Registry (ID: researchregistry5791). Written consents were obtained before participation from the school principal, parents, and participants.

### Participants

A total of 1,296 adolescents (10–14-years-old, 50% girls) from 19 public, subsidised, and private schools of the Valparaiso region (Chile) participated in this study (Table 1). A subsample of 76 participants was recruited voluntarily to be part of the neuroimaging measurements (magnetic resonance imaging (MRI) scan). Exclusion criteria for neuroimaging analysis were: a) no T1 image (n=13), b) incidental tumour finding (n=1), c) no diffusion tensor imaging (DTI) or image quality concern (n=5). Thus a total of 57 right-handed adolescents (26 girls, 47%) from 10 schools were included in the neuroimaging analysis. The power sample estimation for the two parts of this study can be found in our protocol paper ^28^.

**Table 1.** Descriptive participants characteristics.

| Variables | Total sample<br>(n=1,296) | Neuroimaging<br>subsample (n=57) |
| --- | --- | --- |
|  | Mean (SD) / Frequency (%) |  |
| Age (years) | 11.9 (1.2) | 11.6 (1.1) |
| Sex |  |  |
| Girls | 648 (50%) | 31 (54%) |
| Boys | 648 (50%) | 26 (46%) |
| Peak High Velocity (maturation) | -0.41 (1.3) | -0.67 (1.1) |
| Socioeconomic factors |  |  |
| Parent education |  |  |
| None | 869 (67.1%) | 25 (43.9%) |
| Only one | 242 (18.7%) | 16 (28.1%) |
| Both | 185 (14.3%) | 16 (28.1%) |
| School type |  |  |
| Public | 456 (35.2%) | 15 (26.3%) |
| Subsidised | 514 (39.7%) | 26 (45.6%) |
| Private | 326 (25.2%) | 16 (28.1%) |
| School Vulnerability Index |  |  |
| Low | 326 (25.2%) | 16 (28.1%) |
| Middle | 360 (27.8%) | 23 (40.4%) |
| High | 610 (47.1%) | 18 (31.6%) |

Values are displayed as mean or frequency and SD or percentage, respectively, according to data features.

### Socioeconomic factors

Socioeconomic factors are crucial during brain development and have been associated with healthy behaviours, cognitive performance and academic achievements ^17,29–31^. We considered multiple measures of socioeconomic factors. First, we included a measure of parental education level categorised according to the number of parents holding a university degree as a) noneb) one, c) both ^30^. Second, as school characteristics seem to be a stronger predictor of adolescents’ cognitive and school performance in Latin America than socioeconomic status ^32^, we included two school-based indicators. The first was school type (public, subsidised, or private) because of its strong relationship with cognition, health behaviours, and academic performance, and its accuracy as an indicator of socioeconomic and parental education levels in the Chilean context ^33^. The second was school vulnerability index, a Chilean metric that measures the degree of socioeconomic vulnerability of pupils who attend schools with partial or total state funding (subsidised and public schools, respectively) based on the educational level of parents-tutors, student health condition, physical and emotional wellbeing, and school location. This index score ranges from 0 to 100, with a score of zero being assigned to private schools; thus schools are classified as low (<10), middle (≥10 to <60), and high (≥60) ^31^.

### Health-related behaviours measurements

All measurements were carried out in schools, during two four-hour visits per school, separated by eight days. In the first visit, cognitive performance and anthropometry tests were taken, whereas physical fitness was evaluated during the second visit. Cognitive performance and physical fitness were evaluated first in the morning, and questionnaires were administered afterwards.

As part of health-related behaviours, we aimed to measure different lifestyle components either via objective measures or by self-reported questionnaires (Table 2). Body composition was measured by: a) body mass index (BMI), b) waist-to-height ratio (WHtR), and c) four skinfolds. Details on the procedure used to acquire these measurements have been reported elsewhere ^31^. Physical activity (self-reported) was evaluated through two measures, a) active commuting and b) physical activity level. The first was evaluated by a question from the Youth Activity Profile questionnaire (YAP-SL) ^34^, whilst the second measure was acquired via a validated Chilean questionnaire ^35^. Physical fitness was assessed through the ALPHA fitness test battery, which evaluates three main fitness components a) muscular fitness (the maximum handgrip strength test plus the standing long jump test), b) cardiorespiratory fitness, and c) speed/agility fitness ^36^. Each component was then Z-scored and adjusted for age and sex. Diet was evaluated using three indicators (self-reported): a) adherence to the Mediterranean diet ^37^, b) having breakfast on the day that cognitive tests were carried out (see below), and c) quality of breakfast (what ingredients were or were not present, e.g. dairy products, cereals, bread and fruits, and fruit juice). In the case of the last two measures, they were included based on their relationship to cognitive performance in this sample of adolescents ^38^. Finally, sleep problems were quantified via the Spanish version of the Sleep Self-Report questionnaire ^39^.

**Table 2.** Descriptive Health-related behaviours measurements.

|  | Total sample<br>(n=1,296) | Neuroimaging<br>subsample (n=57) |
| --- | --- | --- |
| Variables | Mean (SD) /<br>Frequency (%) |  |
| Body composition |  |  |
| Body Mass Index | 21.5 (3.8) | 21.3 (3.5) |
| Waist-to-Height Ratio | 0.46 (0.1) | 0.45 (0.1) |
| Sum of four skinfolds | 65.1 (27.4) | 61.4 (22.1) |
| Physical activity |  |  |
| Physical activity score | 4.7 (1.3) | 4.8 (1.5) |
| Active commuting |  |  |
| No (none) | 916 (70.7%) | 46 (80.7%) |
| Only one way | 165 (12.7%) | 5 (8.8%) |
| Yes (both) | 215 (16.6%) | 6 (10.5%) |
| Physical fitness components |  |  |
| Muscular fitness |  |  |
| Handgrip strength test (kg) | 23.0 (6.4) | 21.8 (5.7) |
| Standing long jump test (cm) | 141.8 (27.3) | 144.0 (22.3) |
| Cardiorespiratory fitness (min) | 4.0 (2.2) | 4.0 (2.1) |
| Speed and Agility fitness (s) | 13.0 (1.3) | 12.8 (1.2) |
| Diet |  |  |
| Mediterranean Score | 8.6 (2.2) | 9.1 (2.0) |
| Having Breakfast before the cognitive test |  |  |
| No | 367 (28.3%) | 16 (28.1%) |
| Yes | 929 (71.7%) | 41 (71.9%) |
| Breakfast quality score | 2.5 (0.9) | 2.4 (0.8) |
| Sleep problems |  |  |
| Sleep Self-Report Score | 12.4 (5.5) | 10.8 (5.1) |

Values are displayed as mean or frequency and SD or percentage, respectively, according to data features.

### Cognitive and academic achievements

We sought to capture differences in pupils’ cognitive skills and academic achievements. (Table 3). The adolescents’ cognitive performance was evaluated through eight neurocognitive tasks from the NeuroCognitive Performance Test (NCPT) from Lumos Labs, Inc ^40^. This battery included: “Trail Making A and B” assessing attention, cognitive flexibility, and processing speed; the “Forward Memory Span’’ and the “Reverse Memory Span” evaluating short-term visual memory and working memory; the “Go/No-Go” test, assessing inhibitory control and processing speed; the “Balance Scale,” indicating quantitative and analogical reasoning; the “Digit Symbol Coding,” evaluating processing speed; and finally, the “Progressive Matrices,” assessing problem-solving and reasoning/intelligence ^17^. Each test was scaled following a normal inverse transformation of the percentile rank ^40^.

**Table 3.** Descriptive cognitive and academic achievements.

|  | Total sample<br>(n=1,296) | Neuroimaging<br>subsample<br>(n=57) |
| --- | --- | --- |
| Variables | Mean (SD) |  |
| Cognitive performance |  |  |
| Trail making test A (Reverted) | 100.0 (14.7) | 103.6 (15.7) |
| Trail making test B (Reverted) | 100.0 (14.7) | 105.1 (13.2) |
| Forward memory span | 100.0 (14.4) | 101.0 (12.7) |
| Reverse memory span | 100.0 (14.3) | 103.2 (13.1) |
| Go/No-Go (Reverted) | 100.0 (14.7) | 101.1 (15.4) |
| Balance Scale | 100.1 (14.5) | 104.3 (13.5) |
| Digit Symbol Coding | 100.0 (14.7) | 100.3 (15.2) |
| Progressive Matrices | 100.1 (14.3) | 101.2 (13.3) |
| Academic achievement |  |  |
| Language (grades) | 5.40 (0.8) | 5.59 (0.8) |
| Mathematics (grades) | 5.36 (1.0) | 5.59 (0.9) |
| Science score (grades) | 5.48 (0.8) | 5.68 (0.8) |

Values are displayed as mean and SD according to data features.

Academic achievement was established by asking students their general average of the last semester in language, mathematics, and science. In Chile, the grade scoring range is between 1 to 7 points. These three subjects are part of the Programme for International Student Assessment.

### MRI acquisition

All images were obtained with a 1.5 Tesla MRI scanner (AVANTO, Siemens Medical Systems, Erlangen, Germany). Structural MRI: T1 weighted (T1w) three-dimensional rapid gradient echo sequence (3D MPRAGE): TR = 2200 ms; TE = 2,6 ms; flip angle = 8°; FOV = 250 mm; voxel size: 1 × 1 × 1 mm. Sequence duration : 4 min 32 s. Diffusion-weighted MRI (DW-MRI): EPI 2d sequence; b values = 0, 1000 s/mm2, with 20 diffusion-weighted directions; TR = 3300 ms; TE 86ms; voxel size: 1.8 × 1.8 × 5.0 mm; multiband acceleration GRAPPA factor = 2. Sequence duration : 4.02 min. The entire procedure can be reviewed elsewhere ^28^.

### Structural MRI processing

Preprocessing of T1w-MRI data was performed using FSL BET^41^ and the FSL Voxel Based Morphometry (VBM ^42^) pipeline (https://fsl.fmrib.ox.ac.uk/fsl/fslwiki/). Structural images were brain extracted, and tissue segmentation was performed ^41^. After quality control, images were aligned to the MNI152 standard-space T1 template using an optimised combination of linear and non-linear registration (FLIRT followed by FNIRT) ^43,44^. A second quality controlled step ensured there was good alignment to standard space for all subjects. For each subject, we quantified both a voxel-based morphometry feature map of grey-matter (VBM) and a voxel-wise map of the Jacobian deformation (JD). Together, these metrics quantify macroscopic brain differences in grey-matter volume across participants. Finally, these images were concatenated into two 4D images and fed to group-level statistics.

### Diffusion MRI processing

DWI-MRI data was corrected for between-volumes head-movement and eddy currents using FSL Eddy^45^. No reverse phase-encoded image was acquired. Diffusion tensor imaging (DTI) fitting was conducted with FSL DTIFIT^46^. FA (fractional anisotropy), MD (mean diffusivity) and MO (mode of anisotropy) images were then fed into the TBSS pipeline ^46,47^ creating a study-specific, mean FA skeleton, following the procedures used in Salvan et al.,^27^. Together, these metrics quantify brain differences in white-matter microstructure across participants. These images were then fed to group-level statistics.

### Statistical analysis

A study schematic model is presented in **Figure 1**. The first step was to identify modes of covariation between physical and cognitive/academic variables via canonical correlation analysis (CCA). The second step was to characterise independent components of brain structure and microstructure in the sub-sample of pupils with neuroimaging and to test the association with CCA modes. Finally, the third step was to run a mediation analysis to test whether brain components mediate the relationship between physical and cognitive variables.

**Figure 1.**
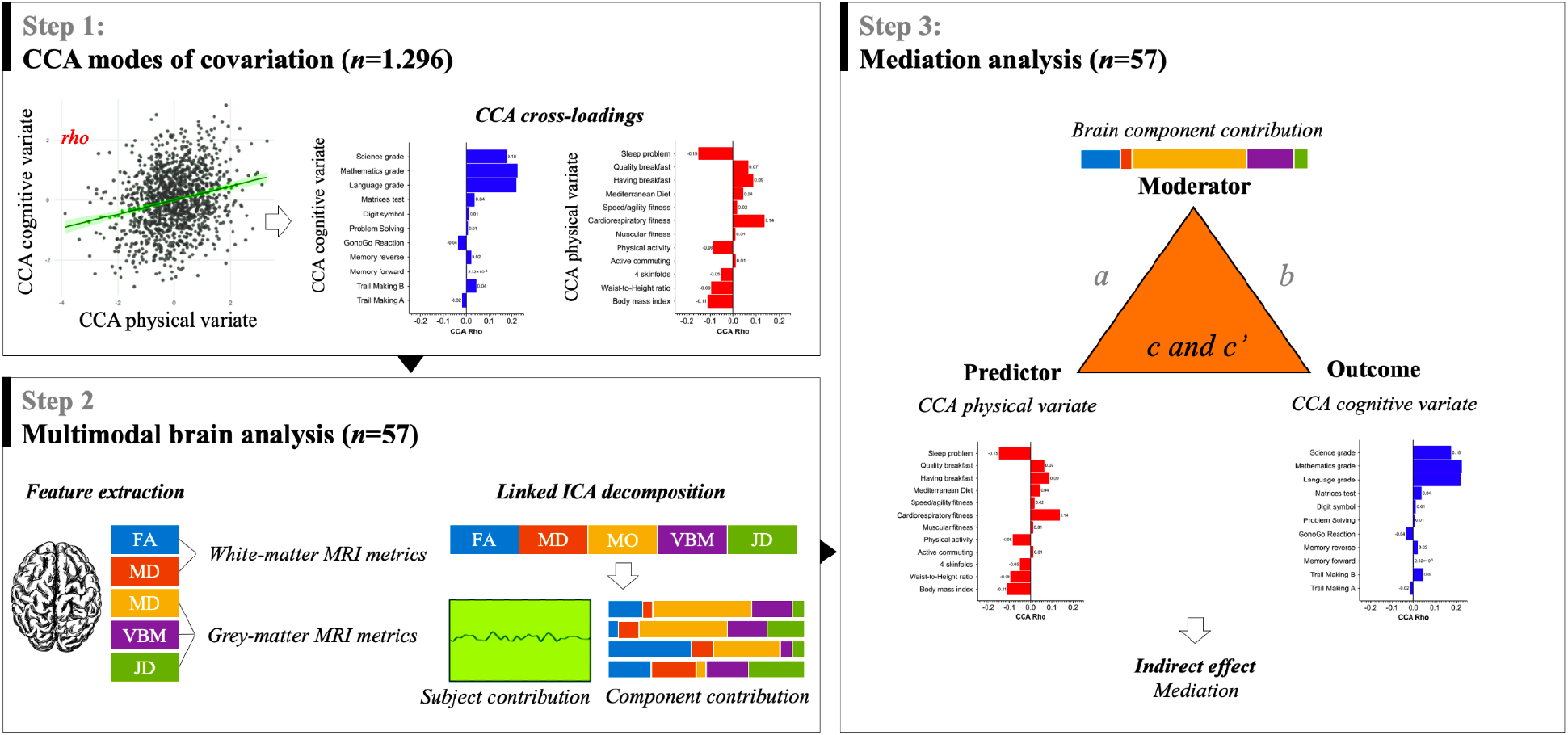
Schematic illustrating the analysis steps. CCA: Canonical correlation analysis; CCA Rho: non-parametric correlation; FA: Fractional anisotropy; MO: Mode of anisotropy; MD: Mean diffusivity; VBM: Voxel-based morphometry; JD: Jacobian deformation; ICA: independent component analysis; a: equation between predictor and moderator; b: equation between moderator and outcome; c: total effect; c’: direct effect.

#### Imputation

Prior to all statistical testing, any missing data were imputed based on the nonparametric missing value method using random forest through the “missForest” R package ^48^. This function successfully imputes large and complex mixed-type datasets (quantitative and/or categorical variables), including complex interactions and non-linear relations by a random forest trained on the observed values predicting the missing values. Prior to imputation, amounts of missing data ranged between 1.2% (i.e., PHV and BMI) to 42.1% (i.e., parental education).

#### Confounds

Seven covariates were included as confound regressors in our analyses. Age, sex, and maturation are relevant factors associated with behaviour and brain development ^2,49^. The differences between chronological and biological age could be reflected in both brain development and cognitive abilities ^50^. Hence, we calculated as a maturity indicator the peak high velocity (PHV), subtracting the PHV age from the chronological age ^51^. In addition, parental education, school type, and school vulnerability index (described above) were included as confounds. Finally, analyses including brain metrics were adjusted for the total brain volume estimated via FreeSurfer (http://surfer.nmr.mgh.harvard.edu/).

#### Canonical correlation analysis (CCA)

Using CCA we sought to characterise modes of covariation relating two sets of variables: a) health-related behaviour measurements (12 variables) and b) cognitive and academic achievements (11 variables). This approach identifies modes of covariation between the two sets of variables making no prior assumptions about relationships given the canonical cross-loading or strength of correlation (CCA rho) that each variable exerted on its opposite canonical variate. Each mode is characterised by a *pair* of CCA canonical variates or CCA subject-vectors, that are maximally correlated. The total number of modes generated is always equal to the number of variables in the smaller dataset (*here* 11). To perform CCA we used the script permcca ^52^ (https://github.com/andersonwinkler/PermCCA) whilst adjusting for confounds of no interest (age, sex, PHV, parental education, school type, and school vulnerability index). The significance of CCA modes was calculated via nonparametric inference testing through 1,000 permutations among subjects within schools (*k*=19), respecting dependencies given by the hierarchical structure of the data ^53^. Family wise error correction (FWE-corr) was applied across all CCA modes in order to correct for multiple comparisons. For those CCA modes deemed significant at FWE-corr p < 0.05, CCA imaging and physical cross-loadings were then extracted.

#### Multimodal neuroimaging networks via linked factorisation

We then aimed to characterise parsimonious multimodal patterns of brain structure and microstructure using the structural and microstructural neuroimaging metrics described above. To do this, we performed a multivariate joint-decomposition called FLICA (FMRIB’s Linked Independent Component Analysis, https://fsl.fmrib.ox.ac.uk/fsl/fslwiki/FLICA) ^42,54^. FLICA is an entirely data-driven approach that can co-model multiple imaging modalities. Its main goal is to model the imaging data as a set of interpretable features (independent components, ICs), most of them characterising biophysically plausible modes of variability across all subjects’ images. Unlike in a principal component analysis, the mixing matrix vectors of an ICA are not forced to be orthogonal to each other and thus can explain the common variance of variables external to the ICA, such as age ^55^. FLICA was implemented as described in detail in earlier papers ^55,56^. Here, we ran FLICA on five different neuroimaging metrics (FA, MD, MO, VBM, and JD) with 15 components. Although previous large-scale studies have performed FLICA with a higher dimensionality (e.g. 484 subjects and 70 ICs) ^55^, here, we chose a smaller number of ICs because of the relatively small sample size of 57 pupils. Hence, we expect to identify coarse structural brain networks with a lower granularity compared with previous studies. To test the robustness of our findings to varying the number of FLICA ICs, we also ran FLICA with 14 and 16 components and repeated all statistical tests.

#### Testing the association between CCA modes and multimodal neuroimaging networks

We next sought to test the association between inter-subject differences in the identified CCA modes (linking health-behaviour with cognitive and academic variables) and differences in multimodal neuroimaging networks in the subset of pupils with neuroimaging data. We tested the regression between the pairs of CCA canonical variates (for each mode: one behavioural and one cognitive) against inter-individual differences in FLICA ICs subject-scores. This was done via a built-in FLICA algorithm ^42,54^ that performs Bonferroni correction across ICs tested and non-imaging measures tested. Thus reported results are fully corrected for multiple comparisons. However, as this test does not take into account the hierarchical structure represented by schools, to corroborate the findings we performed mixed-linear models explicitly modelling the effect of schools (as a random effect).

Furthermore, although the CCA modes were covaried for several confounds of no interest, there is no guarantee that CCA variance in the subset of pupils with neuroimaging data is independent of such effect. Hence we further tested whether such associations were independent of the effect of confounds of no interest (age, PHV, sex, school vulnerability index, parent education, school type, and intracranial volume).

#### Causal mediation analysis

A causal mediation analysis was used to test the mediator role of a brain pattern from the multimodal brain analysis (FLICAs) in the relationship between CCA canonical variates predictor (left CCA variate depicting variance in health-related behaviours) and outcome (right CCA variate depicting variance in cognitive and academic achievements). The analysis was conducted using the R ‘mediation’ function (https://cran.r-project.org). The mediation was based on the Baron-Kenny procedure, and standard errors, confidence intervals, and significance levels were calculated based on a quasi-Bayesian Monte Carlo approximation. Four parameters were estimated the direct effect (path c, non-mediated effect of health-related behaviours on cognitive/academic achievements), b) the indirect effect (path ab, mediated effect of health-related behaviours on cognitive/academic achievements), c) total effect (path c’ = ab + c), and d) the proportion mediated (proportion of the total effect mediated *via* the mediator)^57^.

Because inter-subjects variance in CCA mode 2 (*fitness–cognition mode*) was significantly correlated with school vulnerability index and school type (respectively, Pearson’s rho = 0.35, p<0.01; rho = 0.33, p<0.05; no associations were found for other confounds, see **Figure S6**), causal mediation analysis was performed whilst adjusting for these confounds of no interest.

## Results

### Covariation modes link health-related behaviours to cognitive skills and academic achievements, independent of socioeconomic factors

The primary aim of this study was to characterise modes of covariation linking sets of health-related behaviours and cognitive and academic achievements in a large sample of Chilean adolescents, independent of several confounds of no interest. Using CCA, we found two significant modes of covariation. The first mode links all three academic variables positively with cardiorespiratory fitness and negatively with sleep problems and fatness indicators (CCA rho = 0.29, FWE-corr p-value <0.001; **Figure 2A**). The second mode of covariation links cognitive academic variables positively with all physical fitness components and physical activity (*fitness– cognition mode*; CCA rho = 0.23, FWE-corr p-value <0.001; **Figure 2B**). Importantly, these modes of covariation are independent of age, PHV, parental education, school administration, and school vulnerability index, and they are present after accounting for the cluster effect of school.

**Figure 2.**
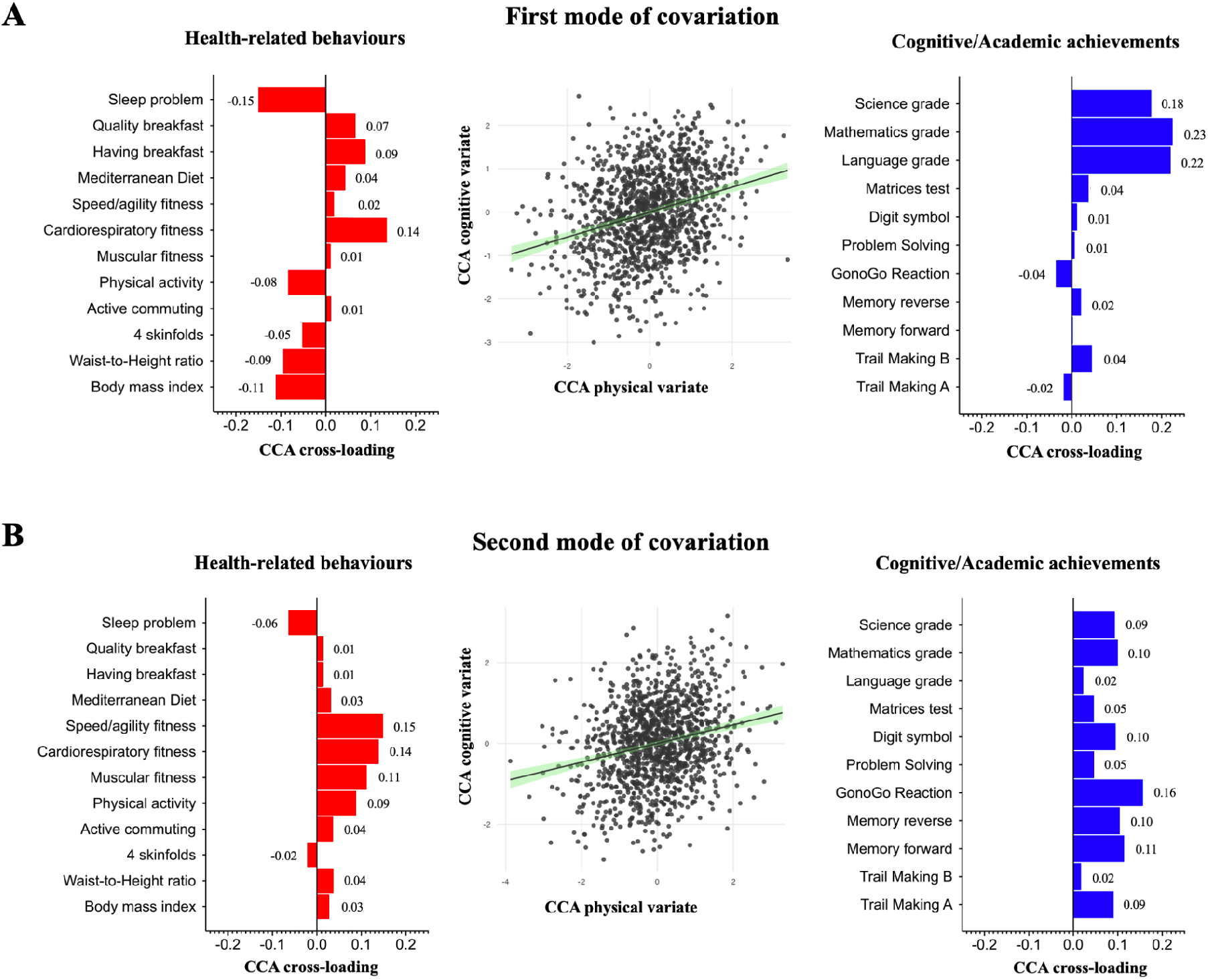
Modes of covariation link sets of health-related behaviours with sets of cognitive/academic variables in adolescents. Showing modes of covariation identified via CCA, adjusted for age, sex, PHV, parental education, school type, school vulnerability index, and school effects. **A**) First mode: pupils with few sleep problems and lower fatness, were those who showed greater academic achievement. **B**) Second mode: pupils with a phenotype of greater physical fitness, were those who also showed greater cognitive skills. Of interest, the two modes of covariation are by construction orthogonal, hence independent from each other. Red: CCA cross-loadings for health-related variables. Blue: CCA cross-loadings for cognitive and academic variables. (*n*=1,296).

### Multimodal brain correlates of fitness–cognitive covariation

The secondary aim was to test whether the identified modes of covariation, linking different sets of health-related behaviours with cognitive/academic achievements, are associated with inter-subjects variation in multimodal MRI patterns of brain structure and microstructure. Using FLICA, we decomposed inter-subjects variation in 5 MRI brain-wide metrics (FA, MD, MO, VBM, and JD) into 15 independent components (FLICA15) or brain networks. Each component is characterised by brain maps of structure and microstructure, depicting the involvement of each MRI metric, and a one-dimensional vector quantifying the subject-wise contribution or, in other words, the subject-scoring along that component. We then used FLICA subject-scorings to test the association between brain structure and microstructure and the identified CCA modes of covariation. After correction for multiple comparisons via a stringent Bonferroni threshold, we found that component IC#10 was significantly associated with both the fitness and cognitive CCA subject-vectors from the second mode of covariation (respectively, p = 0.0067 and p = 0.0026; Bonferroni correction across CCA subject-vectors and FLICA independent components; **Figure 3A and 3B**). This association was robust even after adjusting for confounds of no interest (age, PHV, sex, school vulnerability index, parent education, school type, and intracranial volume) and by using mixed-models accounting for the cluster effect of schools (see Supplementary **Table S4**). All residuals were normally distributed according to Q-Q plots and the Shapiro-Wilk test (result not shown).

**Figure 3.**
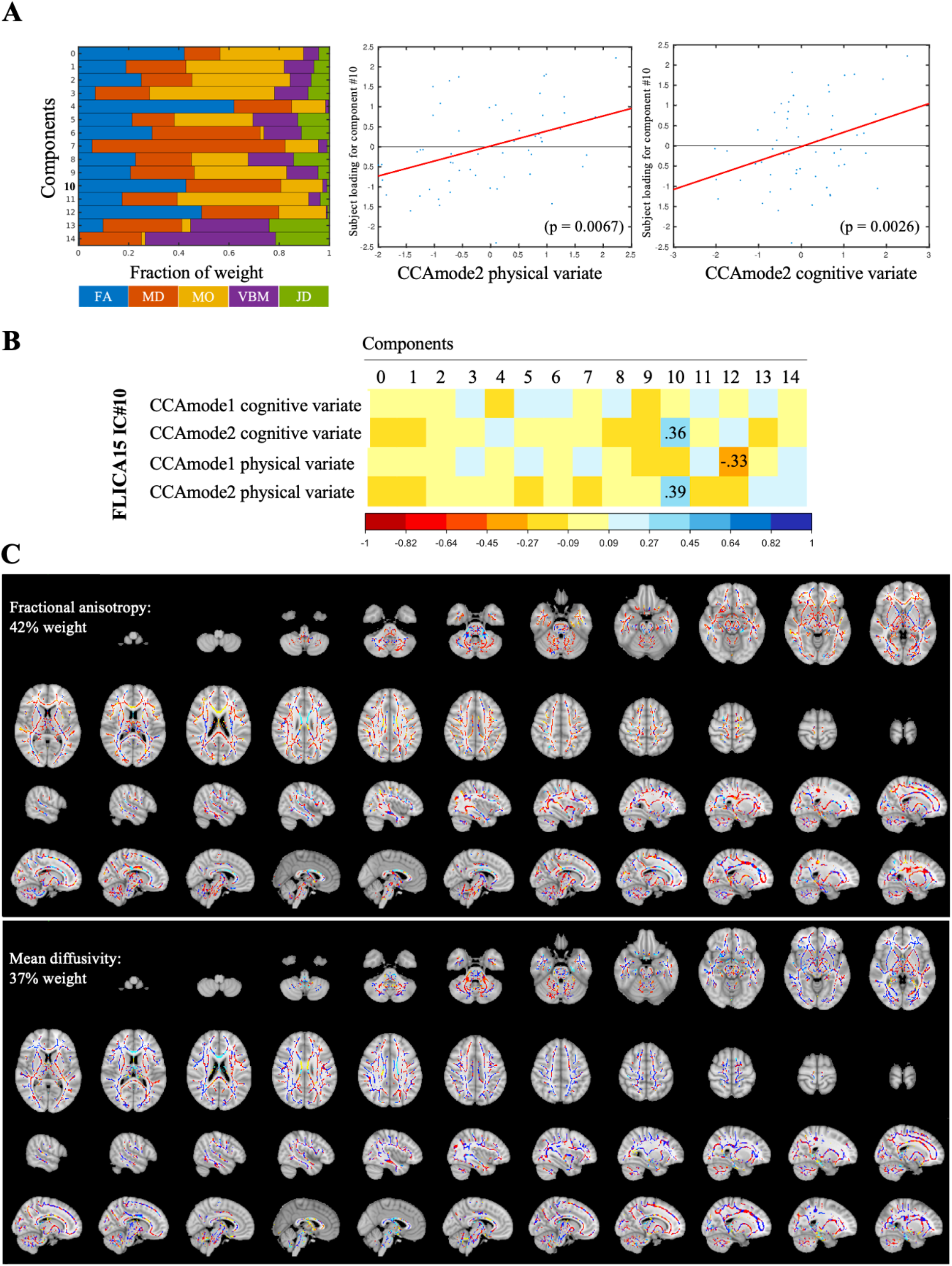
Multimodal neuroimaging correlates of identified covariation modes. Relative weight of brain features in each component. Left: showing involvement of different neuroimaging metrics (FLICA weights). Centre and right showing, the scatter plots of the associations between FLICA15 IC#10 and, respectively, CCA physical and cognitive variates from the second mode of covariation. **B**) Showing correlation coefficients between the two identified modes of covariation and all FLICA15 ICs. Each covariation mode is composed by a pair of CCA variates (see panel **A**). Reporting in numbers only significant associations after multiple comparison correction via Bonferroni–thresholding Only FLICA15 IC#10 was significantly related to both CCA variates for any of the two significant modes. CCACognitive_: CCA cognitive variate. CCAPhysical_: CCA physical variate. **C**) Showing FLICA15 IC#10 brain-wide weights for the two neuroimaging metrics with highest weights (FA and MD).

The brain association with the *fitness-cognition mode* was robust even to changing the number of FLICA components. Indeed both a 14 and 16 factorisation of brain imaging data (respectively, FLICA14 and FLICA16), yielded significant associations with the *fitness-cognition mode* (see **Figure S2**; in FLICA16 the association with the CCA cognitive subject-vectors, although significant per se, did not survive correction for multiple comparisons). The inter-subjects variance of independent components was highly similar to the one characterised by FLICA15 IC#10 (respectively, Pearson’s correlation with FLICA14 IC#8 = 0.89, p-value <0.001; correlation with FLICA16 IC#9 = 0.85, p-value <0.001). Furthermore, the relative contribution of different MRI metrics was also very similar to FLICA15 IC#10 (**Figure 3A**) (FLICA15 IC#10: 42% FA and 37% MD; FLICA14 IC#8: 45% FA and 45% MD; and FLICA16 IC#9: 47% FA and 38%,

**Figures S3 and S4**). Crucially, the spatial maps depicting the independent component coefficients matched the overall modality-specific spatial patterns of FLICA15 IC#10 depicting high coefficients within Corpus callosum FA and MD (**Figures 3C and S5**).

One possibility is that the inter-subject variance in brain microstructure pattern of interest is mainly driven by confounds of no interest (such as sex, age, etc.) or by the cluster effect intrinsic in the sampling (the fact that pupils were sampled across different schools, each potentially representing a cluster with different mean). In order to test whether these factors of no interest drive the variance of the component of interest, we performed voxel-wise non-parametric permutation testing, testing the association between FLICA15 IC#10 and inter-subjects differences in DTI FA whilst controlling for school vulnerability index and school type and whilst constraining permutations allowing shuffling of samples only within the same school. This analysis showed that corpus callosum’s FA was statistically significantly driving the decomposition of FLICA15 IC#10, even after adjusting for socioeconomic indicators (supplementary material, **Figure S5**).

### A trend towards mediation in the fitness–cognitive covariation

Finally, we hypothesised that the identified brain microstructural network (FLICA15 IC#10) could mediate the relationship between fitness and cognitive CCA subject-vectors from the second mode of significant covariation, independent of socioeconomic factors (school vulnerability and school type). As the significance of the association between fitness–brain microstructure (*path a*) and brain microstructure–cognition (*path b*) was previously established (**Figure 3A**), we next performed a causal mediation analysis. We found a trend towards significance for the indirect effect (*path ab*: p-value = 0.088, 95% CI -0.007 to 0.260; **Table 4**) with total effect losing statistical significance without the presence of the mediator (Total effect c: p-value 0.008; Direct effect c’: p-value = 0.092).

**Table 4.** Causal mediation analysis with FLICA15 IC#10 as mediator between CCA co-variates.

|  | Estimate | 95% CI Lower to Upper | p |
| --- | --- | --- | --- |
| Indirect effect | 0.106 | -0.007 to 0.260 | 0.088 |
| Direct Effect | 0.280 | -0.038 to 0.580 | 0.092 |
| Total effect | 0.385 | 0.080 to 0.680 | **0.008** |
| Prop. Mediated | 0.26 | -0.050 to 1.250 | 0.096 |

We further tested the robustness of this trend towards a mediation by replicating the causal mediation analysis for FLICA14 IC#8 and FLICA16 IC#9. FLICA14 showed a significant indirect effect (*path ab*: p-value = 0.010) with the total effect losing statistical significance without the presence of the mediator (Total effect c: p-value 0.010; Direct effect c’: p-value = 0.090). Whilst FLICA16 showed a trend for the indirect path (*path ab*: p-value = 0.076), the total and the direct effects were both statistically significant (p-value = 0.008 and 0.030, respectively) (more details see **Tables S5**).

## Discussion

### Modes of covariation link pupils’ health-related behaviours with cognitive skills and academic achievements

The first aim of this study was to characterise modes of covariation relating sets of health-related behaviours with sets of cognitive and academic achievement variables in a large sample of Chilean adolescents. We found two significant modes of covariation, above and beyond the effect of socioeconomic factors and of other confounds of no interest. The first mode related greater academic achievement with lower sleep problems, higher cardiorespiratory fitness, better diet and body composition markers, and lower levels of physical activity. The second mode related cognitive skills with greater fitness across all physical fitness components tested (cardiorespiratory, muscular, and speed/agility fitness) and with greater physical activity, a phenotype to which we refer to as *physical fitness*.

Our findings are in line with the literature showing an important contribution of multiple healthy-lifestyle behaviours – such as physical activity, physical fitness, sleep hygiene, diet, body composition – in supporting adolescents’ cognitive performance and academic achievements 11,31,58–60. It is indeed well-known that lifestyle behaviours are interdependent, favouring a synergistic beneficial effect on the body ^61^. However, here we further unveil that this lifestyle– cognition covariation is characterised by two distinct and independent latent sources, each characterised by distinct physical and cognitive phenotypes. This result suggests two distinct pathways that could be leveraged via modifiable behaviours in order to improve cognition and academic achievements during adolescence.

Previous studies examining adherence to 24-h movement guidelines ^11,59,60^ (regarding physical activity, sleep, and sedentary behaviour) have found some association between adherence to these guidelines and both cognitive outcomes (4,524 US children aged 8–11 years) ^11^ and academic achievement (1,290 Spanish adolescents aged 11-16 years) ^59,60^. However, there is heterogeneity in these associations depending on the population studied and which combination of guidelines was considered ^59,60^. Therefore, multivariate analyses, integrating more variables reflecting the complexity of human behaviour, might be better suited to identify more robust and plausible modes of variability relating lifestyles and cognition, and hence could provide novel evidence for movement guidelines.

For example, we previously used structural modelling of data from the Chilean Cogni-Action Project,, to test the multivariate association among age, health-related quality of life, school vulnerability index, body mass index, physical activity, and sleep problems with physical fitness and cognitive performance in adolescents ^20^. We found that physical fitness mediated the relationship between school vulnerability index, body mass index, and physical activity and cognitive performance. Using data from the Fit to Study Project in the UK, we previously used CCA to uncover latent factors relating sets of physically active lifestyle measures with mental health and cognition ^26^. The results of the current study further emphasise how modifiable behavioural factors, such as health-related lifestyle, may support cognitive development and academic achievements in adolescence, even in presence of high heterogeneity in the socioeconomic environment.

### Multimodal brain correlates of covariation modes

The second aim of this study was to identify the brain correlates of the identified modes of covariation described above in a subgroup of pupils that underwent neuroimaging. We found that pupils’ differences along the *fitness-cognition mode* were significantly associated with a brain microstructure phenotype. Pupils with both greater cardiovascular fitness and greater cognitive skills were those who also exhibited greater FA and lower MD in the corpus callosum, a pattern of neuroimaging metrics often related to better brain health and cognitive development ^55^. Although this brain network expressed little contribution from brain grey-matter structural metrics, the identified relationship was robust to several confounds of no interest, to the cluster effect due to schools, and to varying the dimensionality of the neuroimaging decomposition method.

There is limited prior literature investigating the multimodal brain correlates of lifestyle behaviours in children and adolescents. Salvan et al. (2021) showed that in a sample of 50 12-year old UK adolescents, a physically active lifestyle (being fitter and less sedentary) was associated with systems-level brain variation across multiple MRI metrics (greater grey-matter perfusion, volume, cortical surface area, greater white-matter extra-neurite density, and resting-state networks activity) ^27^. Although this relationship was robust to a number of confounds of no interest and suggested the presence of diverse biological processes known to be related to change in fitness in rodent experiments, the Authors did not find a significant association with pupils’ cognitive performance. Our findings complement this previous research by showing that a brain-wide pattern of white-matter microstructure relates to interindividual differences in a number of variables related to a healthy-lifestyle as well as to greater cognitive skills.

Traditionally, hippocampal volume and hippocampal connectivity have been associated with higher cardiorespiratory fitness levels in children and adolescents ^62–64^. However several studies have also found beneficial effects of physical training programmes on brain white-matter. For instance, an intervention study reported that children participating in exercise (8-month, 40 minutes of aerobic activities per day) showed improved white-matter microstructure compared to controls ^65^. An after-school program (2h per day for 150 days of moderate-to-vigorous physical activity) found that children who participated in the physical activity program increased white-matter FA and decreased MD in the genu of the corpus callosum ^66^. While another study (8-month, 40 min of aerobic exercise per day) showed that participating in an exercise intervention improves white-matter microstructure in children as compared to a sedentary after-school program ^67^.

Furthermore, previous cross-sectional studies have also reported that physical activity was associated with greater white-matter microstructure ^66^, and cardiorespiratory fitness was associated with greater white-matter volume ^68^, microstructure, and connectivity ^69,70^. Therefore, our findings add to this literature by suggesting that modifiable behaviours such as increasing physical activity and physical fitness could help improve brain development and, in turn, cognitive skills in adolescents.

### Mediation role of brain microstructure

We finally tested whether brain differences mediated the effect of fitness on cognition. Our finding hints at a possible mediator role of healthy brain microstructure on the *fitness–cognition mode* of covariation, independent of socioeconomic status. The association between white-matter microstructure and either executive functions and academic achievements, or active lifestyle markers (i.e., physical activity and physical fitness), both during childhood and adolescence, is well established from previous studies ^71–75^. Several studies have shown a significant association of cardiorespiratory fitness ^69,70^ and physical activity ^66^ with higher FA in sections of the corpus callosum. On the other hand, the corpus callosum is a fundamental white-matter bundle that supports executive functions and academic achievements. Indeed better inhibitory control ^76^, working memory ^77^, task-switching ^78^, language ^79^, and maths performance ^72^ have been linked to higher FA in the corpus callosum. However, very little evidence exists on the mediator role of brain microstructure in the link between cardiorespiratory fitness and cognitive performance.

To the best of our knowledge, only Ruotsalainen, et al., (2020) and Maijer et al., (2021) have explored FA as a moderator or mediator between physical fitness and neurocognitive functioning indicators. Ruotsalainen, et al., (2020) found that in 12-16 years old adolescents the white-matter microstructure of the corpus callosum moderates the association between cardiorespiratory fitness and working memory ^75^. Maijer et al., (2021) found no significant mediation of FA in the relationship between cardiorespiratory fitness and neurocognitive functioning ^80^. Our finding of a trend for a mediating role of the brain phenotype in the relationship between fitness and cognition builds on this existing literature. It hints that greater *physical fitness* may support neurocognitive development by improving brain microstructure in a white-matter bundle that is fundamental for the integration of information across brain networks and that is a key neural correlate of multiple cognitive functions. However, future studies should replicate this mediation analysis with properly powered sample sizes.

### Strengths and limitations

To our knowledge, this is the first study integrating the covariation between health-related behaviours and cognitive/academic variables, with a multimodal neuroimaging analysis in adolescents living in a Latin-American country. These relationships were significant and robust above and beyond the effect of socioeconomic factors, measured via three different variables related to family, school, and social features. This is an important feature of this study that may be of great interest to developing countries and nations with high inequality in income distribution.

While the overall sample in our study was relatively large, our neuroimaging subsample was considerably smaller. In addition, the final sample size for the multimodal neuroimaging analysis had lower statistical power than originally planned ^28^. Therefore, future studies should assess whether the trend for a mediation found in this work can be detected with a bigger sample size.

## Conclusion

The findings reported in this study are of high relevance to public health and educational policies due to the worldwide decline in children and adolescents’ cardiorespiratory fitness ^81^. This is particularly true in countries with high inequality in income distribution ^82^ and low educational achievement, a phenomenon that has been exacerbated globally over the past 15 years ^83^. Our findings show that healthier white-matter microstructure is a clear and robust neural correlate of the relationship between greater cardiorespiratory fitness and higher cognitive performance, independent of effects related to socioeconomic factors. This suggests that promoting health-related lifestyle behaviours could result in a broad brain pattern of microstructural changes that, in turn, may support healthy neurocognitive development in adolescence. Thus, early education focusing on healthy behaviours would be a valuable and low-cost strategy to bridge the cognitive/academic gap due to social inequalities.

## Data Availability

All data produced in the present study are available upon reasonable request to the authors.

## Author Contributions

CC-M, HJ-B, and PS conceived and designed the data analysis and manuscript. CC-M was responsible for coordinating the study, acquiring the data, and writing the first manuscript version. All authors contributed significantly to editing the manuscript and agreed to the final version.

## Data Availability Statement

The data presented in this study are available on request from the corresponding author.

The data are not publicly available as we did not obtain consent for public release of data.

## Acknowledgement

We want to acknowledge Patricio Solis-Urra, Jorge Olivares-Arancibia, Javier Sanchez-Martinez, Tamara Huber-Perez, Steren Chabert and all university students who gathered the data at schools. In addition, we thank school principals, parents, and adolescents who supported this project. We also want to acknowledge the Independent Imagenology Center Quintaimagen, Viña del Mar, Chile. Carlos Cristi-Montero received funding for the Cogni-Action Project from the National Commission for Scientific and Technological Research CONICYT/FONDECYT INICIACION 2016 grant No. 11160703 (Chile), and the National Research and Development Agency (ANID) from Chile-2019, Postdoctoral Grant No. 74200071. The Wellcome Centre for Integrative Neuroimaging is supported by core funding from the Wellcome Trust (203139/Z/16/Z). Heidi Johansen-Berg is supported by a Wellcome Principal Research Fellowship (222446/Z/21/Z). This research was funded in whole, or in part, by the Wellcome Trust (110027/Z/15/Z). For the purpose of Open Access, the author has applied a CC BY public copyright licence to any Author Accepted Manuscript version arising from this submission.

## Supplementary material

**Figure S2.**
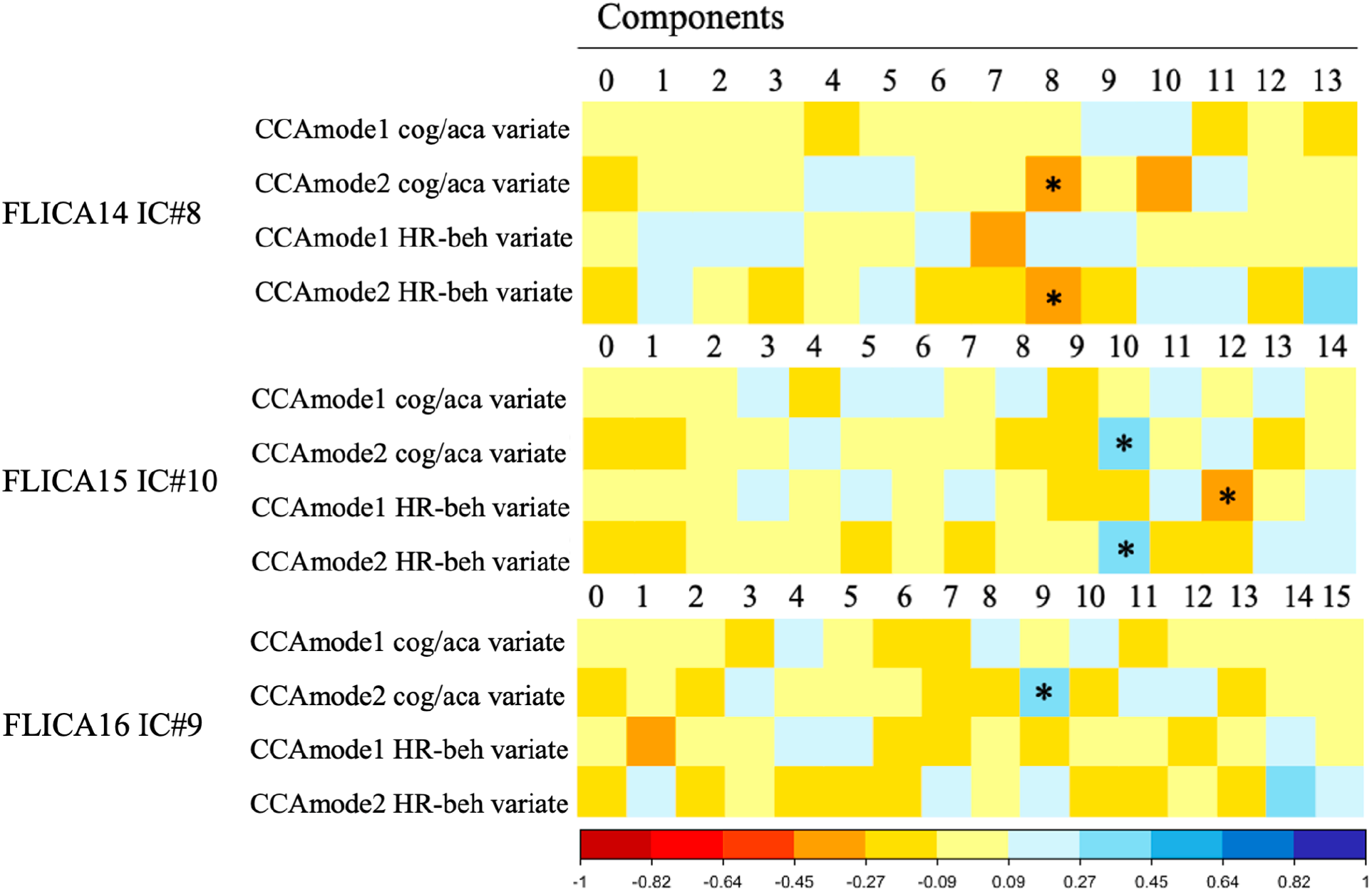
Associations between all four modes of covariations and FLICA14, 15, and 16. Correlation matrix between all four modes of covariation and FLICA’s components. * significant associations (r) after multiple comparisons. The colour indicates the grade of association.

**Figure S3.**
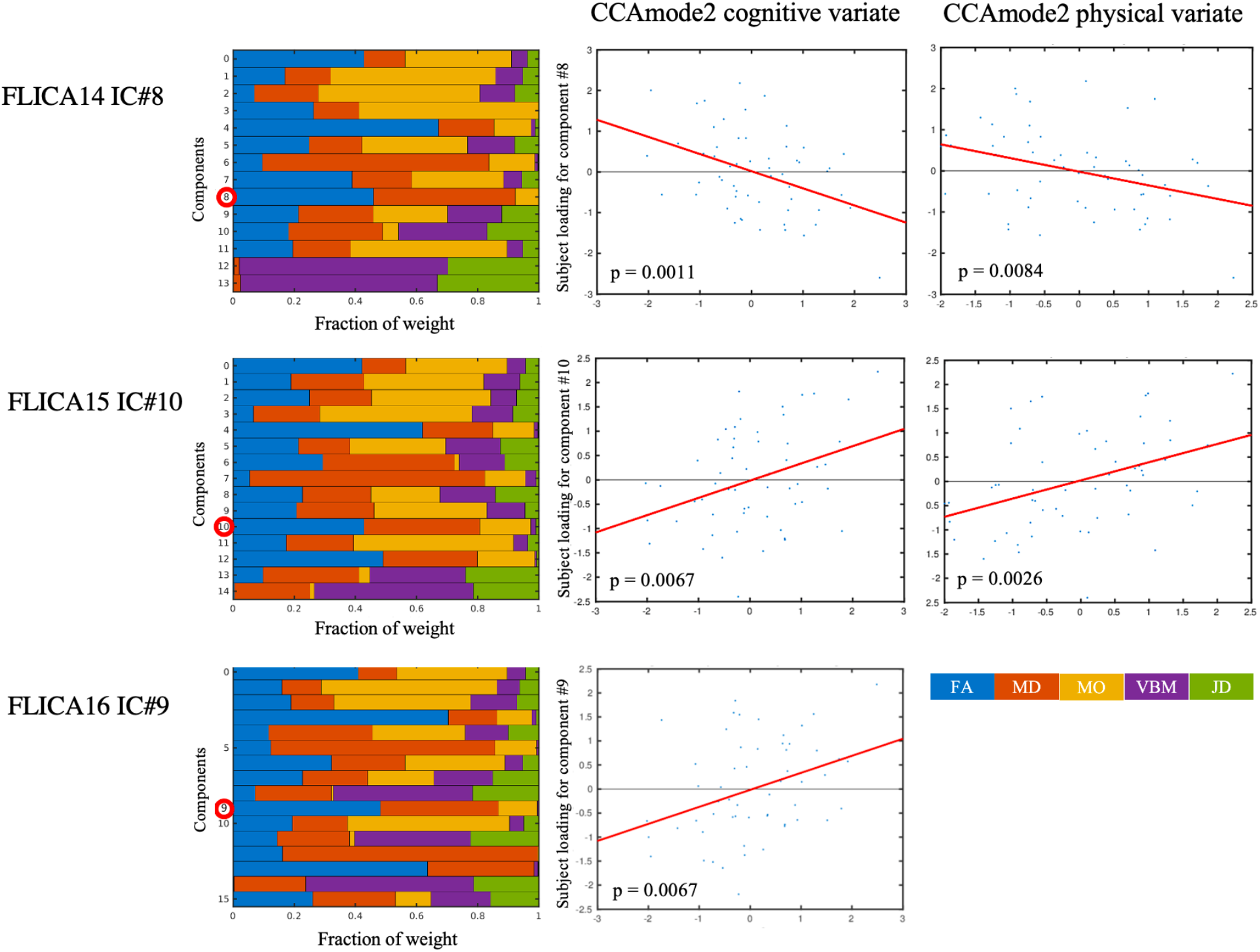
The relative weight of brain features for each component by FLICA and its association with the second mode of covariation. The first column (left) displays the fraction of weight for each component (the red circle represents the component studied). The two columns in the right show the association with the main modes of covariation (cognitive/academic achievements and health-related behaviours, respectively) from the mode of covariation number two. P-value after multiple comparisons.

**Figure S4.**
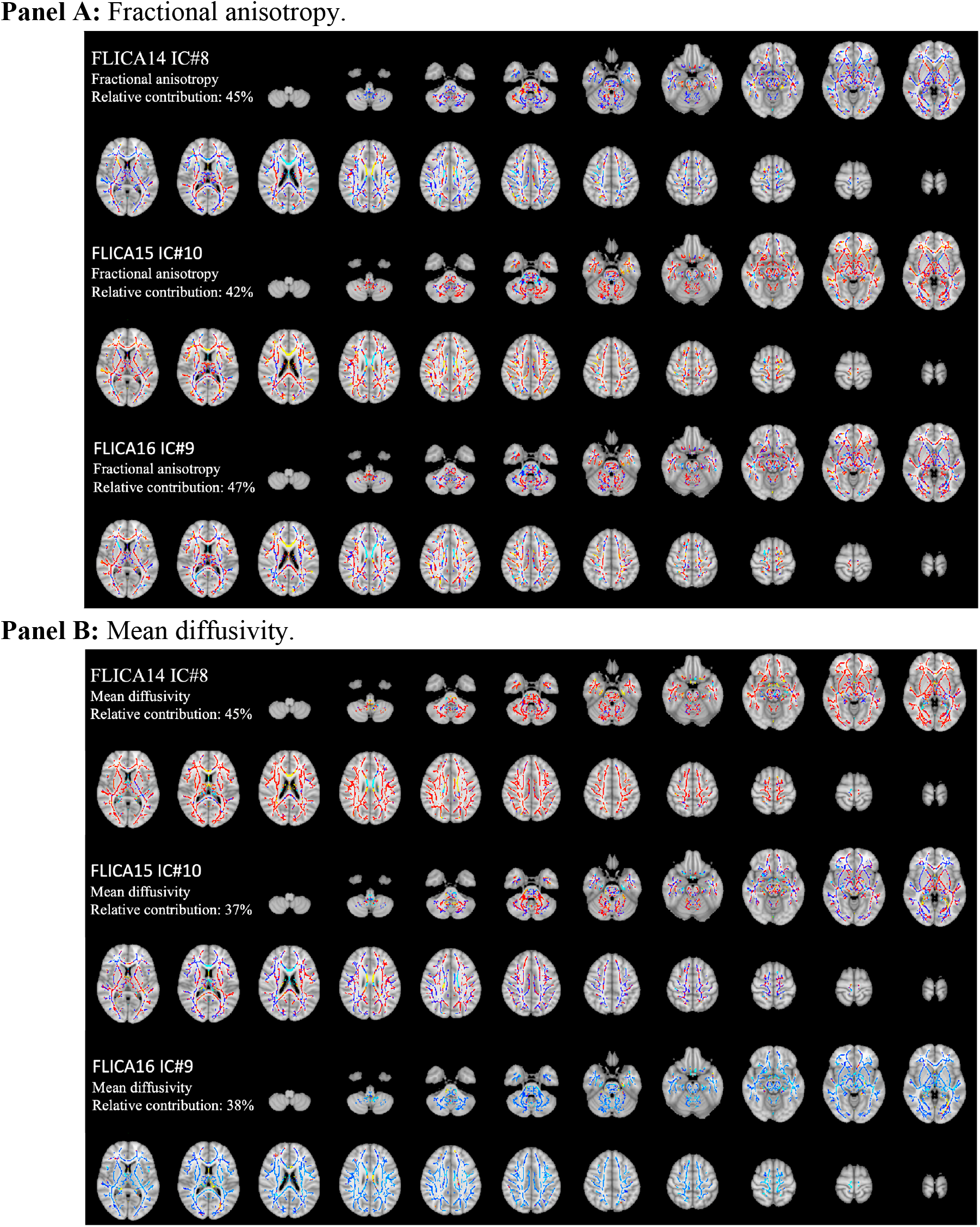
Fractional anisotropy and mean diffusivity relative contributions for main FLICA 14, 15 and 16-components.

**Figure S5.**
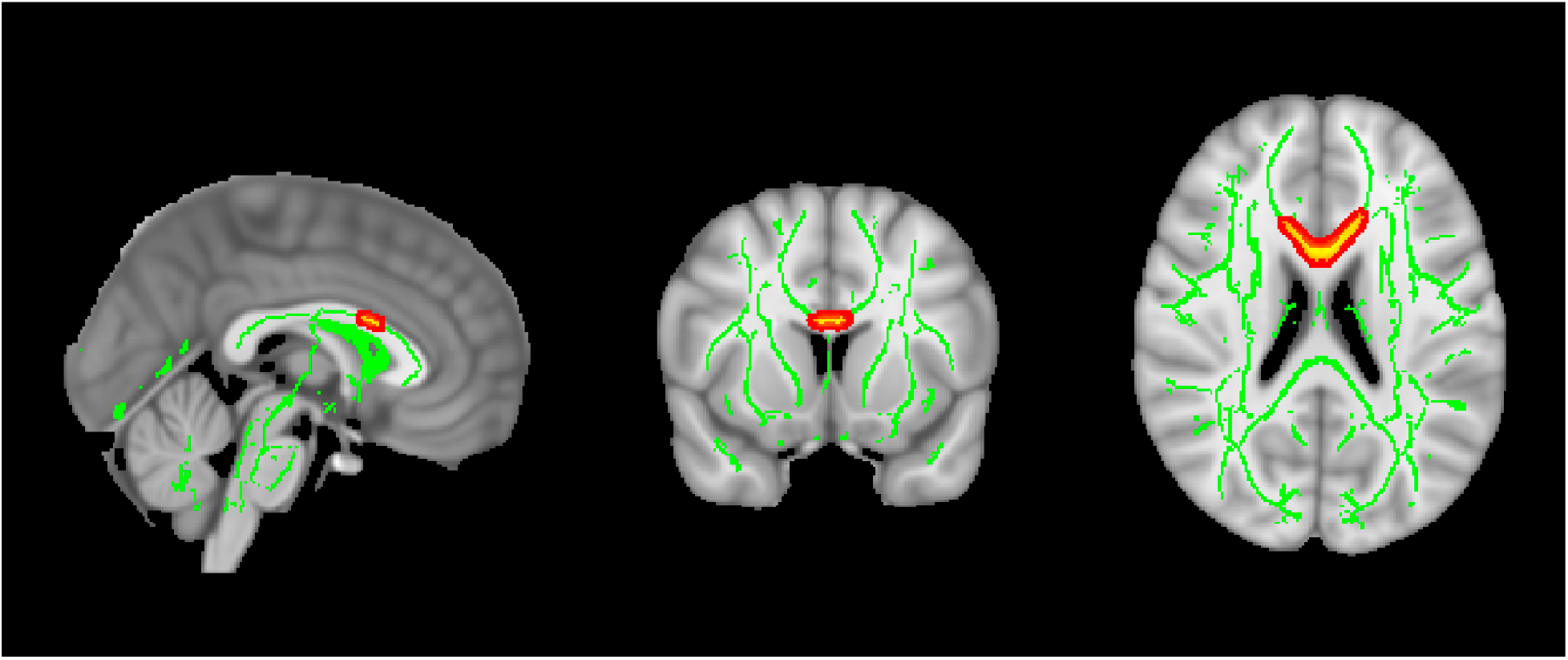
Voxel-wise statistical fractional anisotropy analysis. Figure displays the FA’s tract-based spatial statistics.

**Figure S6.**
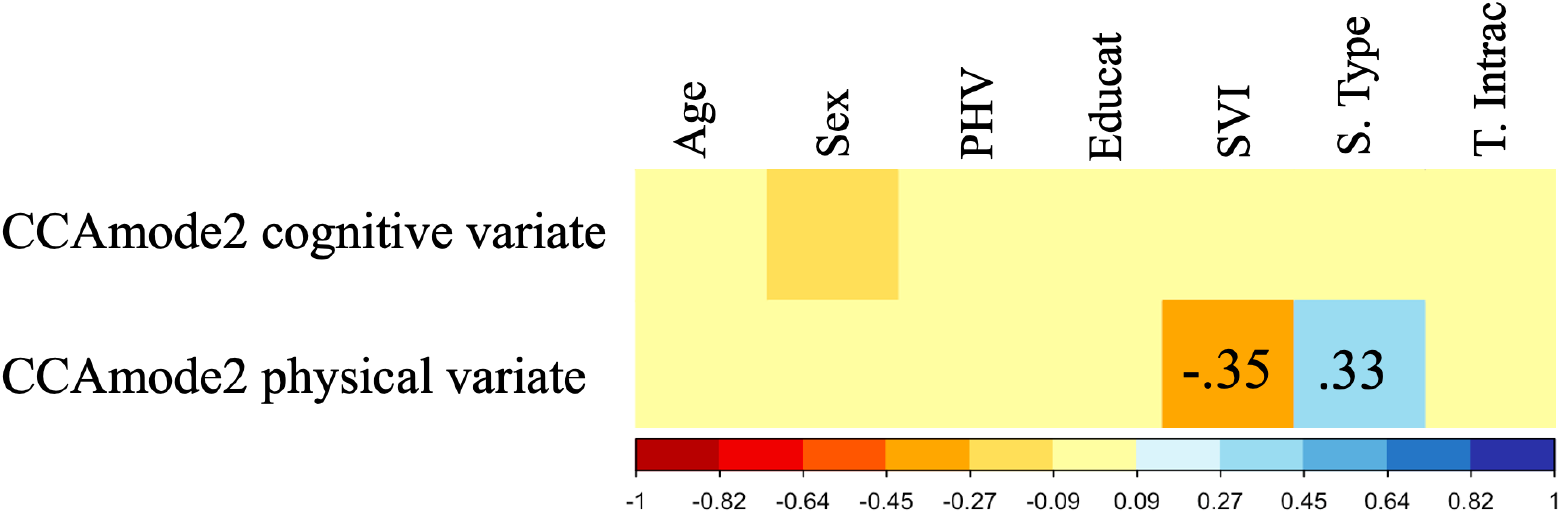
Correlation matrix between the second modes of covariation and all confounds. The plot displays a correlation matrix between the *fitness-cognition mode* and all confounds used in analyses. PHV: peak height velocity (maturation), Educat: Parental education, SVI: school vulnerability index, S. Type: School type (administration), T. Intrac: total intracranial volume.

**Tables S4.**
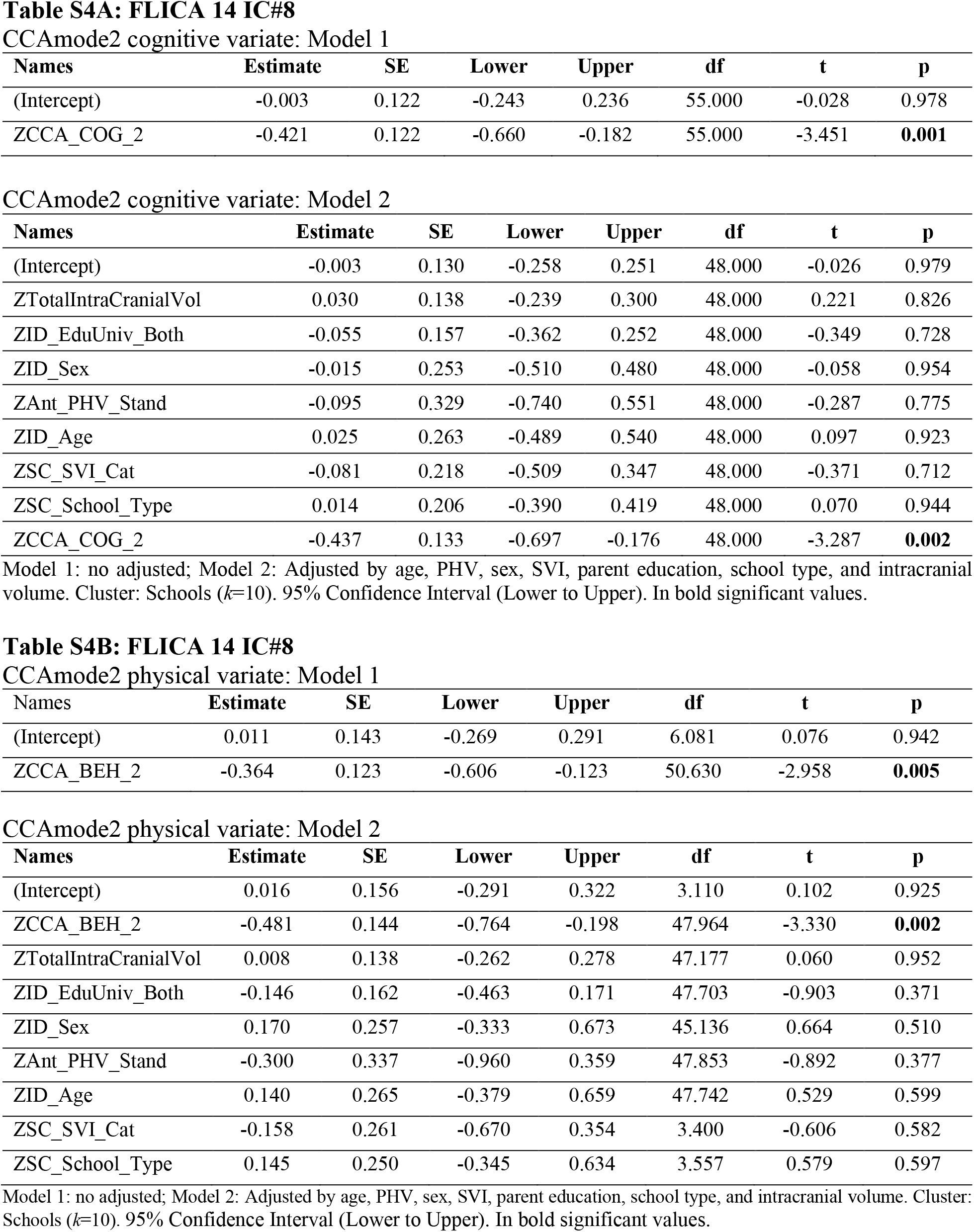
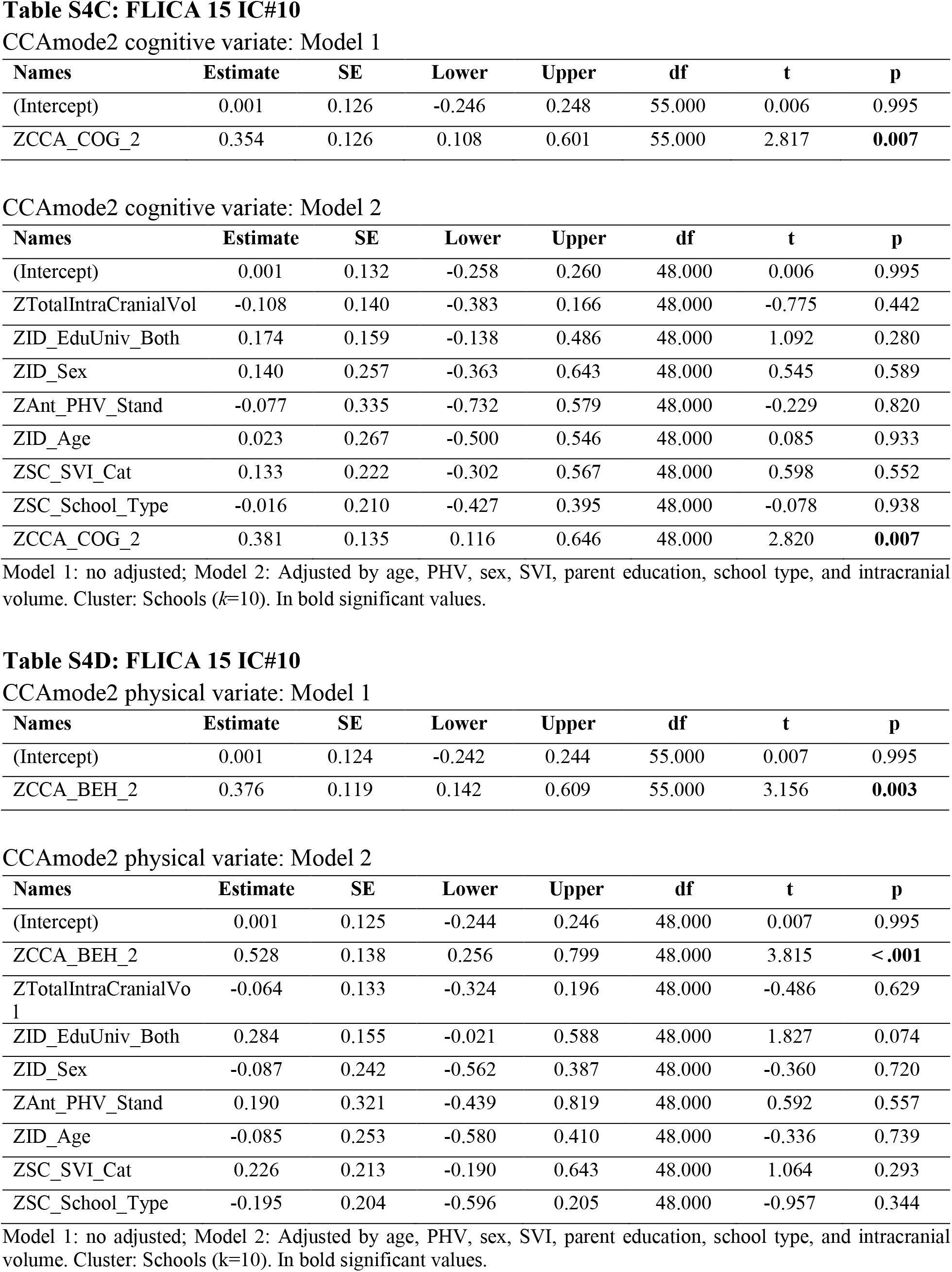
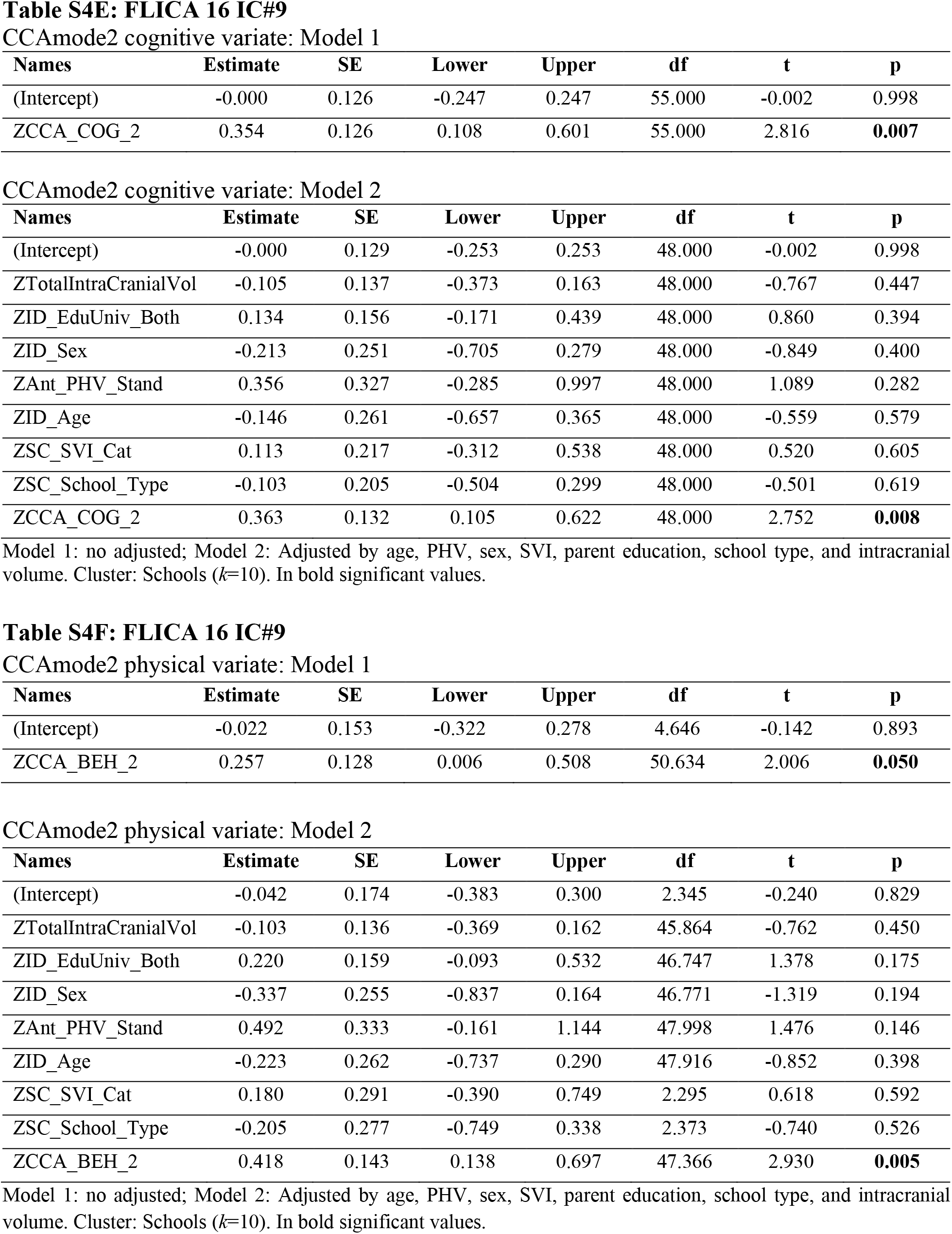
Mixed-models by modes of covariation (fixed effects parameter estimates).

| Names | Estimate | SE | Lower | Upper | df | t | p |
| --- | --- | --- | --- | --- | --- | --- | --- |
| (Intercept) | -0.003 | 0.122 | -0.243 | 0.236 | 55.000 | -0.028 | 0.978 |
| ZCCA_COG_2 | -0.421 | 0.122 | -0.660 | -0.182 | 55.000 | -3.451 | **0.001** |

CCAmode2 cognitive variate: Model 2
| Names | Estimate | SE | Lower | Upper | df | t | p |
| --- | --- | --- | --- | --- | --- | --- | --- |
| (Intercept) | -0.003 | 0.130 | -0.258 | 0.251 | 48.000 | -0.026 | 0.979 |
| ZTotalIntraCranialVol | 0.030 | 0.138 | -0.239 | 0.300 | 48.000 | 0.221 | 0.826 |
| ZID_EduUniv_Both | -0.055 | 0.157 | -0.362 | 0.252 | 48.000 | -0.349 | 0.728 |
| ZID_Sex | -0.015 | 0.253 | -0.510 | 0.480 | 48.000 | -0.058 | 0.954 |
| ZAnt_PHV_Stand | -0.095 | 0.329 | -0.740 | 0.551 | 48.000 | -0.287 | 0.775 |
| ZID_Age | 0.025 | 0.263 | -0.489 | 0.540 | 48.000 | 0.097 | 0.923 |
| ZSC_SVI_Cat | -0.081 | 0.218 | -0.509 | 0.347 | 48.000 | -0.371 | 0.712 |
| ZSC_School_Type | 0.014 | 0.206 | -0.390 | 0.419 | 48.000 | 0.070 | 0.944 |
| ZCCA_COG_2 | -0.437 | 0.133 | -0.697 | -0.176 | 48.000 | -3.287 | **0.002** |

| Names | Estimate | SE | Lower | Upper | df | t | p |
| --- | --- | --- | --- | --- | --- | --- | --- |
| (Intercept) | 0.011 | 0.143 | -0.269 | 0.291 | 6.081 | 0.076 | 0.942 |
| ZCCA_BEH_2 | -0.364 | 0.123 | -0.606 | -0.123 | 50.630 | -2.958 | **0.005** |

CCAmode2 physical variate: Model 2
| Names | Estimate | SE | Lower | Upper | df | t | p |
| --- | --- | --- | --- | --- | --- | --- | --- |
| (Intercept) | 0.016 | 0.156 | -0.291 | 0.322 | 3.110 | 0.102 | 0.925 |
| ZCCA_BEH_2 | -0.481 | 0.144 | -0.764 | -0.198 | 47.964 | -3.330 | **0.002** |
| ZTotalIntraCranialVol | 0.008 | 0.138 | -0.262 | 0.278 | 47.177 | 0.060 | 0.952 |
| ZID_EduUniv_Both | -0.146 | 0.162 | -0.463 | 0.171 | 47.703 | -0.903 | 0.371 |
| ZID_Sex | 0.170 | 0.257 | -0.333 | 0.673 | 45.136 | 0.664 | 0.510 |
| ZAnt_PHV_Stand | -0.300 | 0.337 | -0.960 | 0.359 | 47.853 | -0.892 | 0.377 |
| ZID_Age | 0.140 | 0.265 | -0.379 | 0.659 | 47.742 | 0.529 | 0.599 |
| ZSC_SVI_Cat | -0.158 | 0.261 | -0.670 | 0.354 | 3.400 | -0.606 | 0.582 |
| ZSC_School_Type | 0.145 | 0.250 | -0.345 | 0.634 | 3.557 | 0.579 | 0.597 |
| (Intercept) | 0.001 | 0.126 | -0.246 | 0.248 | 55.000 | 0.006 | 0.995 |
| ZCCA_COG_2 | 0.354 | 0.126 | 0.108 | 0.601 | 55.000 | 2.817 | **0.007** |

CCAmode2 cognitive variate: Model 2
| Names | Estimate | SE | Lower | Upper | df | t | p |
| --- | --- | --- | --- | --- | --- | --- | --- |
| (Intercept) | 0.001 | 0.132 | -0.258 | 0.260 | 48.000 | 0.006 | 0.995 |
| ZTotalIntraCranialVol | -0.108 | 0.140 | -0.383 | 0.166 | 48.000 | -0.775 | 0.442 |
| ZID_EduUniv_Both | 0.174 | 0.159 | -0.138 | 0.486 | 48.000 | 1.092 | 0.280 |
| ZID_Sex | 0.140 | 0.257 | -0.363 | 0.643 | 48.000 | 0.545 | 0.589 |
| ZAnt_PHV_Stand | -0.077 | 0.335 | -0.732 | 0.579 | 48.000 | -0.229 | 0.820 |
| ZID_Age | 0.023 | 0.267 | -0.500 | 0.546 | 48.000 | 0.085 | 0.933 |
| ZSC_SVI_Cat | 0.133 | 0.222 | -0.302 | 0.567 | 48.000 | 0.598 | 0.552 |
| ZSC_School_Type | -0.016 | 0.210 | -0.427 | 0.395 | 48.000 | -0.078 | 0.938 |
| ZCCA_COG_2 | 0.381 | 0.135 | 0.116 | 0.646 | 48.000 | 2.820 | **0.007** |

| Names | Estimate | SE | Lower | Upper | df | t | p |
| --- | --- | --- | --- | --- | --- | --- | --- |
| (Intercept) | 0.001 | 0.124 | -0.242 | 0.244 | 55.000 | 0.007 | 0.995 |
| ZCCA_BEH_2 | 0.376 | 0.119 | 0.142 | 0.609 | 55.000 | 3.156 | **0.003** |

CCAmode2 physical variate: Model 2
| Names | Estimate | SE | Lower | Upper | df | t | p |
| --- | --- | --- | --- | --- | --- | --- | --- |
| (Intercept) | 0.001 | 0.125 | -0.244 | 0.246 | 48.000 | 0.007 | 0.995 |
| ZCCA_BEH_2 | 0.528 | 0.138 | 0.256 | 0.799 | 48.000 | 3.815 | **<.001** |
| ZTotalIntraCranialVol | -0.064 | 0.133 | -0.324 | 0.196 | 48.000 | -0.486 | 0.629 |
| ZID_EduUniv_Both | 0.284 | 0.155 | -0.021 | 0.588 | 48.000 | 1.827 | 0.074 |
| ZID_Sex | -0.087 | 0.242 | -0.562 | 0.387 | 48.000 | -0.360 | 0.720 |
| ZAnt_PHV_Stand | 0.190 | 0.321 | -0.439 | 0.819 | 48.000 | 0.592 | 0.557 |
| ZID_Age | -0.085 | 0.253 | -0.580 | 0.410 | 48.000 | -0.336 | 0.739 |
| ZSC_SVI_Cat | 0.226 | 0.213 | -0.190 | 0.643 | 48.000 | 1.064 | 0.293 |
| ZSC_School_Type | -0.195 | 0.204 | -0.596 | 0.205 | 48.000 | -0.957 | 0.344 |
| (Intercept) | -0.000 | 0.126 | -0.247 | 0.247 | 55.000 | -0.002 | 0.998 |
| ZCCA_COG_2 | 0.354 | 0.126 | 0.108 | 0.601 | 55.000 | 2.816 | **0.007** |

CCAmode2 cognitive variate: Model 2
| Names | Estimate | SE | Lower | Upper | df | t | p |
| --- | --- | --- | --- | --- | --- | --- | --- |
| (Intercept) | -0.000 | 0.129 | -0.253 | 0.253 | 48.000 | -0.002 | 0.998 |
| ZTotalIntraCranialVol | -0.105 | 0.137 | -0.373 | 0.163 | 48.000 | -0.767 | 0.447 |
| ZID_EduUniv_Both | 0.134 | 0.156 | -0.171 | 0.439 | 48.000 | 0.860 | 0.394 |
| ZID_Sex | -0.213 | 0.251 | -0.705 | 0.279 | 48.000 | -0.849 | 0.400 |
| ZAnt_PHV_Stand | 0.356 | 0.327 | -0.285 | 0.997 | 48.000 | 1.089 | 0.282 |
| ZID_Age | -0.146 | 0.261 | -0.657 | 0.365 | 48.000 | -0.559 | 0.579 |
| ZSC_SVI_Cat | 0.113 | 0.217 | -0.312 | 0.538 | 48.000 | 0.520 | 0.605 |
| ZSC_School_Type | -0.103 | 0.205 | -0.504 | 0.299 | 48.000 | -0.501 | 0.619 |
| ZCCA_COG_2 | 0.363 | 0.132 | 0.105 | 0.622 | 48.000 | 2.752 | **0.008** |

| Names | Estimate | SE | Lower | Upper | df | t | p |
| --- | --- | --- | --- | --- | --- | --- | --- |
| (Intercept) | -0.022 | 0.153 | -0.322 | 0.278 | 4.646 | -0.142 | 0.893 |
| ZCCA_BEH_2 | 0.257 | 0.128 | 0.006 | 0.508 | 50.634 | 2.006 | **0.050** |

CCAmode2 physical variate: Model 2
| Names | Estimate | SE | Lower | Upper | df | t | p |
| --- | --- | --- | --- | --- | --- | --- | --- |
| (Intercept) | -0.042 | 0.174 | -0.383 | 0.300 | 2.345 | -0.240 | 0.829 |
| ZTotalIntraCranialVol | -0.103 | 0.136 | -0.369 | 0.162 | 45.864 | -0.762 | 0.450 |
| ZID_EduUniv_Both | 0.220 | 0.159 | -0.093 | 0.532 | 46.747 | 1.378 | 0.175 |
| ZID_Sex | -0.337 | 0.255 | -0.837 | 0.164 | 46.771 | -1.319 | 0.194 |
| ZAnt_PHV_Stand | 0.492 | 0.333 | -0.161 | 1.144 | 47.998 | 1.476 | 0.146 |
| ZID_Age | -0.223 | 0.262 | -0.737 | 0.290 | 47.916 | -0.852 | 0.398 |
| ZSC_SVI_Cat | 0.180 | 0.291 | -0.390 | 0.749 | 2.295 | 0.618 | 0.592 |
| ZSC_School_Type | -0.205 | 0.277 | -0.749 | 0.338 | 2.373 | -0.740 | 0.526 |
| ZCCA_BEH_2 | 0.418 | 0.143 | 0.138 | 0.697 | 47.366 | 2.930 | **0.005** |

Model 1: no adjusted; Model 2: Adjusted by age, PHV, sex, SVI, parent education, school type, and intracranial volume. Cluster: Schools ( $k=10$ ). In bold significant values.

**Table S5A.** Mediation analysis for FLICA 14 IC#8.

|  | **Estimate** | **95% CI Lower to Upper** | **p** |
| --- | --- | --- | --- |
| Indirect effect | 0.126 | 0.022 to 0.280 | **0.010** |
| Direct Effect | 0.260 | -0.033 to 0.540 | 0.090 |
| Total effect | 0.386 | 0.086 to 0.680 | **0.010** |
| Prop. Mediated | 0.31 | 0.050 to 1.210 | **0.020** |

In bold significant values. Sample size: 57, Quasi-Bayesian Confidence Intervals, Simulations: 1000.

**Table S5B.** Mediation analysis for FLICA 16 IC#9.

|  | **Estimate** | **95% CI Lower to Upper** | **p** |
| --- | --- | --- | --- |
| Indirect effect | 0.075 | -0.004 to 0.200 | 0.076 |
| Direct Effect | 0.309 | 0.024 to 0.580 | **0.030** |
| Total effect | 0.385 | 0.094 to 0.680 | **0.008** |
| Prop. Mediated | 0.18 | -0.012 to 0.810 | 0.076 |

In bold significant values. Sample size: 57, Quasi-Bayesian Confidence Intervals, Simulations: 1000.

STROBE Statement–Checklist of items that should be included in reports of ***cross-sectional studies***

In bold significant values. Sample size: 57, Quasi-Bayesian Confidence Intervals, Simulations: 1000. STROBE Statement—Checklist of items that should be included in reports of *cross-sectional studies*
|  | Item No | Recommendation | Page No |
| --- | --- | --- | --- |
| Title and abstract | 1 | (a) Indicate the study's design with a commonly used term in the title or the abstract | 1 |
|  |  | (b) Provide in the abstract an informative and balanced summary of what was done and what was found | 1 |
| Introduction |  |  |  |
| Background/rationale | 2 | Explain the scientific background and rationale for the investigation being reported | 2 |
| Objectives | 3 | State specific objectives, including any prespecified hypotheses | 3 |
| Methods |  |  |  |
| Study design | 4 | Present key elements of study design early in the paper | 3-6 |
| Setting | 5 | Describe the setting, locations, and relevant dates, including periods of recruitment, exposure, follow-up, and data collection | 3 |
| Participants | 6 | (a) Give the eligibility criteria, and the sources and methods of selection of participants | 3 |
| Variables | 7 | Clearly define all outcomes, exposures, predictors, potential confounders, and effect modifiers. Give diagnostic criteria, if applicable | 3-6 |
| Data sources/<br>measurement | 8* | For each variable of interest, give sources of data and details of methods of assessment (measurement). Describe comparability of assessment methods if there is more than one group | -- |
| Bias | 9 | Describe any efforts to address potential sources of bias | 9, 18 |
| Study size | 10 | Explain how the study size was arrived at | 3 |
| Quantitative variables | 11 | Explain how quantitative variables were handled in the analyses. If applicable, describe which groupings were chosen and why | 3-6 |
| Statistical methods | 12 | (a) Describe all statistical methods, including those used to control for confounding | 9 |
|  |  | (b) Describe any methods used to examine subgroups and interactions | -- |
|  |  | (c) Explain how missing data were addressed | 9 |
|  |  | (d) If applicable, describe analytical methods taking account of sampling strategy | -- |
|  |  | (e) Describe any sensitivity analyses | 11 |
| Results |  |  |  |
| Participants | 13* | (a) Report numbers of individuals at each stage of study—eg numbers potentially eligible, examined for eligibility, confirmed eligible, included in the study, completing follow-up, and analysed | 3 |
|  |  | (b) Give reasons for non-participation at each stage | 3 |
|  |  | (c) Consider use of a flow diagram | -- |
| Descriptive data | 14* | (a) Give characteristics of study participants (eg demographic, clinical, social) and information on exposures and potential confounders | 4 |
|  |  | (b) Indicate number of participants with missing data for each variable of interest | 9 |
| Outcome data | 15* | Report numbers of outcome events or summary measures | 11-15 |
| Main results | 16 | (a) Give unadjusted estimates and, if applicable, confounder-adjusted estimates and their precision (eg, 95% confidence interval). Make clear which confounders were adjusted for and why they were included | 29-31 |
|  |  | (b) Report category boundaries when continuous variables were categorized | 4 |
|  |  | (c) If relevant, consider translating estimates of relative risk into absolute risk for a meaningful time period | -- |
| Other analyses | 17 | Report other analyses done—eg analyses of subgroups and interactions, and sensitivity analyses | 25-32 |
| **Discussion** |  |  |  |
| Key results | 18 | Summarise key results with reference to study objectives | 15-18 |
| Limitations | 19 | Discuss limitations of the study, taking into account sources of potential bias or imprecision. Discuss both direction and magnitude of any potential bias | 18 |
| Interpretation | 20 | Give a cautious overall interpretation of results considering objectives, limitations, multiplicity of analyses, results from similar studies, and other relevant evidence | 15-18 |
| Generalisability | 21 | Discuss the generalisability (external validity) of the study results | 15-18 |
| **Other information** |  |  |  |
| Funding | 22 | Give the source of funding and the role of the funders for the present study and, if applicable, for the original study on which the present article is based | 24 |

\*Give information separately for exposed and unexposed groups.
**Note:** An Explanation and Elaboration article discusses each checklist item and gives methodological background and published examples of transparent reporting. The STROBE checklist is best used in conjunction with this article (freely available on the Web sites of PLoS Medicine at http://www.plosmedicine.org/, Annals of Internal Medicine at http://www.annals.org/, and Epidemiology at http://www.epidem.com/). Information on the STROBE Initiative is available at www.strobe-statement.org.

